# Clustering analysis based on PiCCO parameters in patients with septic shock

**DOI:** 10.1101/2022.02.02.22270316

**Authors:** Yanfei Shen, Caibao Hu, Guolong Cai, Jing Yan

**Affiliations:** Department of Intensive Care, Zhejiang Hospital, Hangzhou, Zhejiang, 310000, China

**Author notes:** Corresponding author: Jing Yan., Department of Intensive Care, Zhejiang Hospital, Hangzhou, Zhejiang, China. co-first authors (Yanfei Shen, Caibao Hu).

**Keywords:** acute kidney injury, hemodynamics, mortality, PiCCO, septic shock

## Abstract

The current study identified three septic shock phenotypes according to the pulse index continuous cardiac output parameters: Phenotype-1: normal cardiac output and vascular resistance, high blood volume; Phenotype-2: low cardiac output, normal blood volume, and high vascular resistance. Phenotype-3: normal vascular resistance and blood volume, high cardiac output. The mortality was significantly high in phenotype-1, but comparable between phenotype-2 and phenotype-3 (13/17 (76.4) vs. 13/35 (37.1) vs. 16/46 (34.7), p <0.001). Compared to phenotype 1 and 3, phenotype 2 had a higher AKI incidence and higher creatinine level at discharge.

## Main text

Accurate assessment of hemodynamic status is essential in septic shock, as inappropriate fluid resuscitation may lead to poor outcomes. Although much effort has been made, the optimal balance within cardiac output, blood volume, and vascular resistance remains unclear[1]. For instance, when should inotropic agents be used in resuscitation? Would patients benefit from early vasopressor use through reducing the fluid amount? Several large-scale septic shock trials [2] have investigated the optimal protocols in fluid resuscitation. However, clinical parameters such as mean artery pressure (MAP), central venous oxygenation, and lactate cannot accurately reflect specific hemodynamic status. Whether patients with different hemodynamic status would benefit from different resuscitation strategies and have different prognosis remains unclear.

The pulse index continuous cardiac output (PiCCO) system can more accurately reflect the hemodynamic status, compared to clinical parameters. The current study explored latent septic shock phenotypes based on PiCCO parameters and evaluated whether the prognosis differed within these phenotypes.

Data were extracted from the Medical Information Mart for Intensive Care IV database [3]. The patient’s informed consent was waived due to the retrospective nature. Septic shock patients with PiCCO records were screened. Data on PiCCO measurement, MAP within three hours before and after the first PiCCO measurement, and laboratory indexes were extracted. Missing data of PiCCO parameters were not imputed for accuracy.

Five PiCCO parameters, including cardiac stroke volume index (SVI), cardiac output index (CI), extravascular lung water index (ELWI), global end-diastolic volume index (GEDI), and system vascular resistance index (SVRI), were used for clustering analysis. Non-normal variables were log-transformed and scaled. K-means clustering with Euclid distance was performed using R software[4]. The optimal number of clusters was determined using the elbow method[5].

Ninety-eight septic shock patients with PiCCO records were included. Three phenotypes were identified according to initial PiCCO parameters (Figure 1). Compared to phenotype-1 (Table 1), phenotype-2 had lower CI (3.1 ± 1.1 vs. 2.3 ± 0.6, p =0.025) and GEDI (1067.9 ± 415.5 vs. 711.1 ± 191.8, p <0.001) but higher SVRI (1530.8 ± 715.1 vs. 2617.7 ± 997.2, p <0.001), while phenotype-3 had higher CI (3.1 ± 1.1 vs. 4.0 ± 1.0, p <0.001), lower GEDI (1067.9 ± 415.5 vs. 749.8 ±212.4, p <0.001), and comparable SVRI (1530.8 ± 715.1 vs. 1258.5 ± 357.2, p =0.538). The in-hospital mortality was significantly higher in phenotype-1 than that in phenotype-2 and phenotype-3 (13/17 (76.4) vs. 13/35 (37.1) vs. 16/46 (34.7), p <0.001). Compared to phenotype-2, phenotype-3 had higher CI (2.3 ± 0.6 vs. 4.0 ± 1.0, p <0.001) and lower SVRI (2617.7 ± 997.2 vs. 1258.5 ± 357.2, p <0.001), while the GEDI (711.1 ± 191.8 vs. 749.8 ±212.4, p =1.000) and the in-hospital mortality (13/35 (37.1) vs. 16/46 (34.7), p =0.826) were comparable between phenotype-2 and phenotype-3. Although the hemodynamic status differed within three phenotypes, the MAP status within three hours before and after the initial PiCCO measurement was comparable.

**Table 1.** Comparisons of clinical features within three phenotypes based on PiCCO parameters

|  | Phenotype-1<br>(n = 17) | Phenotype-2<br>(n = 35) | Phenotype-3<br>(n = 46) | P value |
| --- | --- | --- | --- | --- |
| **Initial PiCCO parameters used for clustering analysis** |  |  |  |  |
| *Initial SVI (ml/m<sup>2</sup>)* | 31.8 ± 14.1 | 23.4 ± 8.7* | 45.3 ± 10.1**## | < 0.001 |
| *Initial CI (L/min/m<sup>2</sup>)* | 3.1 ± 1.1 | 2.3 ± 0.6* | **4.0 ± 1.0\*\*##** | < 0.001 |
| *Initial ELWI (ml/kg)* | 26.8 ± 7.2 | 11.3 ± 3.7** | 10.8 ± 3.6** | < 0.001 |
| *Initial GEDI (ml/m<sup>2</sup>)* | **1067.9 ± 415.5** | 711.1 ± 191.8** | 749.8 ± 212.4** | < 0.001 |
| *Initial SVRI (DS\*m<sup>2</sup>/cm<sup>5</sup>)* | 1530.8 ± 715.1 | **2617.7 ± 997.2\*\*** | 1258.5 ± 357.2## | < 0.001 |
| Mean SVI (ml/m<sup>2</sup>) | 33.7 ± 11.5 | 34.6 ± 12.3 | 45.6 ± 9.1**## | < 0.001 |
| Mean CI (L/min/m<sup>2</sup>) | 3.1 ± 0.9 | 3.1 ± 1.0 | 3.9 ± 0.7**## | < 0.001 |
| Mean ELWI (ml/kg) | 28.0 ± 18.5 | 15.9 ± 19.3 | 18.7 ± 23.9 | 0.167 |
| Mean GEDI (ml/m<sup>2</sup>) | 896.8 ± 157.3 | 749.8 ± 179.6* | 767.2 ± 198.0* | 0.022 |
| Mean SVRI (DS*m<sup>2</sup>/cm<sup>5</sup>) | 1751.9 ± 958.2 | 1932.3 ± 565.2 | 1458.8 ± 419.1## | 0.002 |
| **MAP within three hours before and after the first PiCCO measurement** |  |  |  |  |
| Initial MAP | 72.2 ± 15.0 | 73.6 ± 15.8 | 74.8 ± 16.6 | 0.855 |
| Maximum MAP | 87.0 ± 14.2 | 91.8 ± 14.8 | 91.8 ± 31.5 | 0.760 |
| Minimum MAP | 61.5 ± 9.6 | 59.4 ± 21.2 | 64.9 ± 19.4 | 0.428 |
| Mean MAP | 73.0 ± 9.1 | 78.2 ± 12.7 | 77.7 ± 18.3 | 0.470 |
| **Laboratory markers & disease scores** |  |  |  |  |
| Initial creatinine | 1.5 ± 0.9 | 1.5 ± 0.8 | 1.5 ± 1.6 | 1.000 |
| Maximum creatinine | 2.3 ± 1.2 | 3.1 ± 2.4 | 2.4 ± 2.0 | 0.233 |
| Last creatinine before discharge | 0.7 ± 0.2 (n = 4) | 1.5 ± 1.3 (n = 22) | 1.0 ± 0.9 (n = 30) | 0.217 |
| Initial WBC | 10.3 ± 8.7 | 14.8 ± 15.2 | 13.1 ± 7.0 | 0.379 |
| Initial P/F | 138.3 ± 184.3 | 313.9 ± 218.4* | 297.9 ± 210.3* | 0.025 |
| Initial lactate | 5.7 ± 4.2 | 4.4 ± 2.9 | 3.4 ± 2.7* | 0.039 |
| Initial SOFA | 5.1 ± 3.5 | 3.9 ± 3.1 | 2.6 ± 3.5* | 0.039 |
| Maximum SOFA | 14.7 ± 3.2 | 13.0 ± 2.9 | 12.0 ± 4.5* | 0.047 |
| **Clinical outcomes** |  |  |  |  |
| ICU LOS | 2.9 (1.7 – 12.9) | 11.2 (6.1 – 18.6)* | 12.6 (6.8 – 19.2)** | 0.044 |
| Hospital LOS | 2.8 (1.5 – 15.9) | 19.1 (9.3 – 30.9)* | 19.8 (12.4 – 27.3)* | 0.025 |
| **AKI (all)** | 12 (70.5) | 31 (88.5) | 34 (73.9) | 0.191 |
| **AKI (grade 3)** | 5 (29.4) | 14 (40.0) | 11 (23.9) | 0.296 |
| **In-hospital death** | 13 (76.4) | 13 (37.1)** | 16 (34.7)** | 0.009 |

Abbreviations: PiCCO pulse index continuous cardiac output; SVI stroke volume index; CI cardiac output index; ELWI extravascular lung water index; GEDI global end-diastolic volume index; SVRI system vascular resistance index; MAP mean artery pressure; WBC white blood cell; P/F oxygen partial pressure/fraction of inspired oxygen ratio; SOFA sequential organ failure assessment; LOS length of stay; AKI acute kidney injury.
Note: \* and \*\* represented p < 0.05 and p < 0.01, respectively, compared to phenotype-1. # and ## represented p < 0.05 and p < 0.01, respectively, compared to phenotype-2.

**Figure 1.**
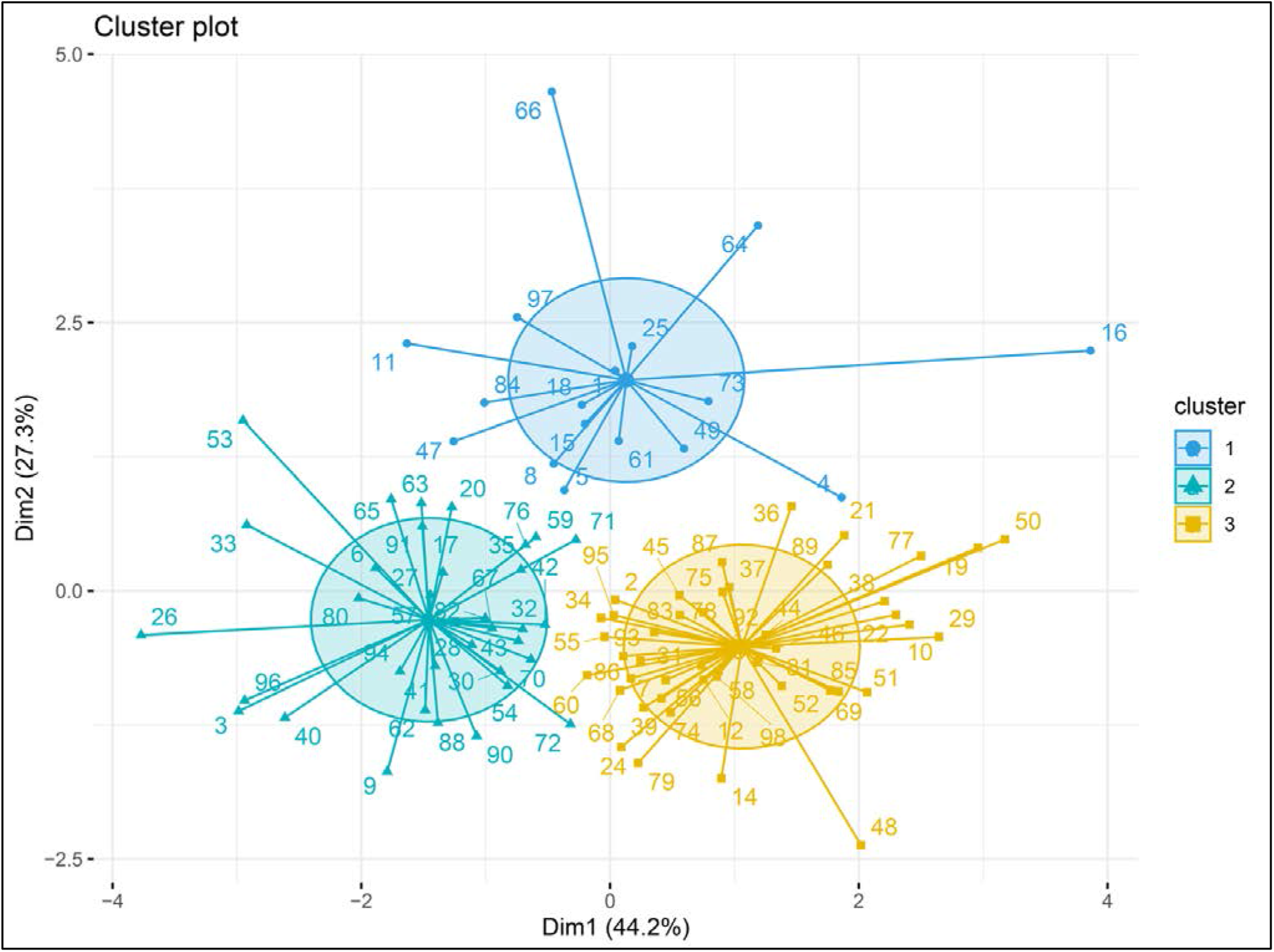
Three phenotypes were identified in clustering analysis. Circles, triangles and squares represented individual patients.

According to our findings, similar hemodynamic goals (MAP) can be achieved by high vascular resistance or high cardiac output. The mortality was comparable between these two phenotypes (2 and 3). However, patients with excessive fluid retention had higher mortality than those with high vascular resistance or cardiac output. Therefore, it will be particularly interesting to determine whether there is a benefit in using multiple therapies in early resuscitation (such as early use of vasopressors and inotropic agents) given concerns about excessive fluid administration. In addition, compared to phenotype 1 and 3, phenotype 2 had a higher AKI incidence and higher creatinine level at hospital discharge. Although the comparison was not statistically significant (small sample size), it is worth investigating whether high vascular resistance is associated with potential renal injury.

The limitation of the current study is that only an association, not causation, was established between hemodynamic status and mortality/AKI. Whether the prognosis would be improved by hemodynamic status-guided interventions still needs further investigation.

## Data Availability

All data produced are available online at https://mimic.mit.edu/.

## Declaration

### Ethics approval and consent to participate

Not applicable.

### Consent for publication

Not applicable.

### Availability of data and material

Not applicable.

### Authors’ contributions

Y.S. and J.Y. came up with the question. Y.S and C.H. was responsible for data analysis and G.C. was responsible for writing. All authors reviewed the manuscript.

## Acknowledgements

Not applicable.

## Reference

1. Lat I, Coopersmith CM, De Backer D, Research Committee of the Surviving Sepsis C, Members of the Surviving Sepsis Campaign Research Committee contributing to this article are as f, Co-chair AGA, Co-chair BB, Manhasset NY, Consultant SWA, Barcelona S et al: The Surviving Sepsis Campaign: Fluid Resuscitation and Vasopressor Therapy Research Priorities in Adult Patients. Critical care medicine 2021, 49(4):623–635.

2. Investigators P, Rowan KM, Angus DC, Bailey M, Barnato AE, Bellomo R, Gimbel E et al: Early, Goal-Directed Therapy for Septic Shock - A Patient-Level Meta-Analysis. The New England journal of medicine 2017, 376(23):2223–2234.

3. Johnson AE, Pollard TJ, Shen L, Lehman LW, Feng M, Ghassemi M, et al: MIMIC-III, a freely accessible critical care database. Scientific data 2016, 3:160035.

4. Seymour CW, Kennedy JN, Wang S, Chang CH, Elliott CF, Xu Z, et al: Derivation, Validation, and Potential Treatment Implications of Novel Clinical Phenotypes for Sepsis. JAMA 2019, 321(20):2003–2017.

5. Wilkerson MD, Hayes DN: ConsensusClusterPlus: a class discovery tool with confidence assessments and item tracking. Bioinformatics 2010, 26(12):1572–1573.

